# Gene expression differences in differentially methylated sites associated with HIV status and cocaine use

**DOI:** 10.1101/2024.11.03.24316634

**Authors:** Eric J. Earley, Bryan C. Quach, Fang Fang, Laura J. Bierut, M-J S. Milloy, Kanna Hayashi, Kora DeBeck, Dana B. Hancock, Bradley E. Aouizerat, Ke Xu, Eric Otto Johnson

## Abstract

**Background:** Epigenome studies of human HIV-1 (HIV) in whole blood have uncovered a growing list of differentially methylated genes associated with either HIV acquisition, disease progression, or both. Cocaine use is associated with increased disease severity, and methylation changes in some of the HIV-associated genes mediate this effect. Many of these genes are critical players in innate immune response, including both regulators and targets of interferon-alpha and NF-kB activation. However, no study to date has evaluated the gene expression dynamics for these genes in the context of HIV.

**Methods:** Targeted gene expression analyses were performed on 588 people who used illicit drugs within a harmonized cohort comprised of the Vancouver People Who Inject Drugs Study (VPWIDS) using RNAseq in whole blood, including 227 people living with HIV (PLWH). Eighteen genes were selected from six recent epigenome-wide association studies to test for differential expression by HIV status. Both gene-level and transcript-level expression changes were estimated using negative binomial regression models.

**Results:** Nine of the 18 target genes exhibited significant upregulation in PLWH after multiple hypothesis testing correction: *EPSTI1, IFI44L, IFIT3, MX1, NLRC5, PARP9, PLSCR1, RIN2*, and *RSAD2*. Transcript-level analysis detected additional upregulation of isoforms for genes *CD44, RASSF3*, and *TAP1*. Stratified analysis by cocaine use revealed *MX1* and *RSAD2* to be exclusively upregulated among PLWH who recently used cocaine. Pathway analysis identified significant dysregulation in the interferon alpha/beta signaling pathway.

**Conclusions:** We confirm the dysregulation of genes previously reported to have differential methylation among PLWH. Results from this study support the model of epigenetic changes altering gene expression for key immune genes such as *NLRC5* and *MX1*, and demonstrate systemic dysregulation of genes involved in innate immune function.

## INTRODUCTION

Epigenetic changes are associated with human immunodeficiency virus-1 (HIV-1) integration into the human genome, as well as viral latency and disease severity.^1-7^ DNA methylation (5mC), the most commonly studied epigenetic modification, regulates gene expression by covalently bonding a methyl group to cytosine-guanine dinucleotides (CpG) and preventing transcription factor binding^8^. While advances in antiretroviral therapy (ART) have made HIV a chronic and manageable disease for those with adequate access and exposure to treatment, infection is still linked to increased risk of non-AIDS defining conditions such as cancer and impaired cognitive function.^7^ Methylation and gene expression dynamics appear to play a role in these comorbid disease-related process.^9-13^

Recent epigenome-wide association studies (EWAS) have uncovered a growing list of differentially methylated CpGs near genes that regulate cellular immunity and response to infection ^7,14-19^. Multiple studies have shown hypomethylation of the promoter regions of genes *NLRC5*^*14-18*^ and *MX1*^*15,18*^ and hypermethylation of *CX3CR1*^*15,18*^ and *TNF*^*17*^ in people living with HIV (PLWH). These genes are key players in cellular immunity, stress response, inflammation, NF-kB signaling cascade, and activation of MHC class I genes.^20-23^ Developing a more complete model of gene expression dynamics in PLWH may uncover novel therapeutic targets and reduce the risk of non-AIDS defining complications.

The use of cocaine, a prevalent unregulated drug among PLWH, is associated with accelerated HIV progression.^24-27^ In one longitudinal study, PLWH who report using crack cocaine at least daily exhibited higher disease severity independent of exposure to ART.^28^ Molecular studies show the biological markers of HIV disease severity such as CD4^+^ cell counts and viral load are worse in the context of cocaine use.^24,25,29^ Recent studies suggest heightened severity is partially mediated by DNA methylation, potentially through alterations in DNAm of immune response genes and subsequent changes in cytokine gene expression.^30,31^

Despite replicated studies showing consistent association of CpGs with HIV acquisition or severity, none of the genes proximal to the published CpG sites have been tested for changes in gene expression. In the current study, we tested gene expression of genes proximal to the CpG sites associated with HIV acquisition, disease severity, or both. Expression was measured in people who inject drugs (PWID) from the Vancouver People Who Inject Drugs Study (VPWIDS).

## METHODS

### Cohort description

Details of the VPWIDS have been previously described^32^. Briefly, VPWIDS participants originated from either new community recruitment or selection from existing cohort studies. For the new recruitment, participants were recruited from Vancouver’s Downtown Eastside neighborhood, an area with widespread illicit drug use and HIV infection. Participants were ≥ 18 years old, used any illicit drug via injection at least once in the 30 days prior to enrollment, and provided written informed consent. Recruited participants completed an interviewer-administered questionnaire to provide harmonious data with existing studies on socio-demographics, substance use patterns, and sexual behaviors; underwent a test of HIV serostatus; and if HIV seropositive, plasma HIV RNA-1 viral load and viral genotyping.

For the selection from existing cohort studies, VPWIDS participants were drawn from either (1) the AIDS Care Cohort to evaluate Exposure to Survival Services (ACCESS), a prospective cohort of people living with HIV infection who use illicit drugs; or (2) the Vancouver Injection Drug Users Study (VIDUS), a prospective cohort of people at risk of HIV infection who use illicit drugs. VIDUS/ACCESS participants completed an interviewer-administered questionnaire on socio-demographics, HIV risk behaviors, HIV treatment patterns, substance use patterns, social/structural exposures, and other relevant exposures/outcomes.

The VIDUS, ACCESS and VPWIDS studies were reviewed and approved by the University of British Columbia/Providence Healthcare research ethics board. All VPWIDS participants provided blood specimens for extraction of human DNA and mRNA extraction. All participants were of European descent. Recent cocaine use was defined as reporting cocaine use at least once in six months before the baseline interview. Viral suppression was defined as <200 viral copies/mL of whole blood. ^33^

### Sex as a biological variable

Our study enrolled self-reported male and female participants, although we note the majority of participants were male. To reduce the risk of accidental mis-labeling of bio-samples during DNA and RNA sequencing, discrepancies between self-reported sex and chromosomally defined sex resulted in exclusion of that participant in the study.

### Deriving target genes from HIV-associated CpGs

A literature search for EWAS within HIV acquisition, severity, or both, was performed using PubMed by searching for keyword terms “HIV” and “epigenome.” To ensure cross-study comparability, citations were excluded unless data were derived *in vivo* from human whole blood. In addition, only epigenome-wide studies were considered. Studies focused on interrogating the epigenetic clock were excluded. Six publications were selected by this process, and from these 18 CpG sites were selected. A list of proximal protein-coding genes to these CpG sites was identified, and these 18 genes were subject to differential gene and transcript expression analysis (**Supplemental Table 1**).

### RNA sequencing and DNA Genotyping

RNAseq libraries were prepared from whole blood samples using the NuGEN Universal Plus mRNA-seq kit with human globin AnyDeplete on the Illumina Nextseq platform by the Rutgers University Cell and DNA Repository (RUCDR Infinite Biologics). Trimmed reads were mapped to the GRCh38 human reference transcriptome (GENCODE v28) with HISAT2 v2.1.0.^34^ Transcript-level quantification of RNA was estimated with Salmon v0.11.2 ^35^ using GC bias correction, and gene-level quantifications were derived from these transcript quantification estimates using the R library, *tximport*.^36^ Additional quality control metrics were generated using FASTQC.^37^

VPWIDS participants’ DNA was genotyped on the Illumina Infinium OmniExpress-24 BeadChip. DNA variant calling was also performed on samples from the same study participants and concordance between RNA and DNA single nucleotide variants (SNVs) and small insertions-deletions (indels) was used to identify potentially mislabeled samples. RNA variants were called using the *mpileup* function within *SAMtools*, ^38^ and problematic sample pairs between genotyping array and RNAseq were identified using Pearson’s *r* correlation. If samples with conflicting IDs exhibited *r* >0.8 between DNA and RNA variant calling they were assumed to be incorrectly labeled and IDs were updated to match each other. If samples with matching IDs exhibited *r* >0.6 they were assumed to be correctly matched. Discrepancy between the *k*-means (*k*=2) clustering of sex assignments and the mean Y chromosome gene expression identified a further set of potentially problematic samples. As a final QC step, we excluded samples with low transcript diversity (lower quartile - 1.5*IQR), high read duplication (>75%), and missing genotype data. Overall, these filters removed 14 samples resulting in a final analysis set of 588 samples, 227 HIV+ samples (i.e., samples from PLWH), and 361 HIV-samples (i.e., samples from people without HIV).

### Cell type deconvolution

To control observed gene expression bias due to variation in cell types across individuals, we performed cell type deconvolution using the tool CIBERSORTx ^39^ with the LM22 leukocyte cell-type reference.^40^ All 22 cell types were used to estimate per-sample cell type proportions. However, to replicate the approach of prior methylation studies,^14-18^ only 5 cell classes were considered in differential expression models – CD4+ T cells, CD8+ T cells, B cells, Granulocytes, and Monocytes. The higher resolution proportions of the 22 cell types were collapsed into these categories via summing the sub-types (e.g., resting Mast cells + activated Mast cells + Eosinophils + Neutrophils = Granulocytes) except for NK T cells which were not present in the LM22 reference.

### Statistical analysis

Unless otherwise noted, all reported *P*-values were adjusted for multiple hypothesis testing using the conservative Bonferroni correction to reduce type 1 error rates. Cohort characteristics were tested for association with HIV status in univariate logistic regression. Differential expression analysis was performed with *DESeq2*,^41^ which fits negative binomial regression models for each gene/transcript with the gene/transcript expression as the outcome variable and HIV status as the primary independent variable. Age, sex, RNA quality (RIN), 5 deconvolution-derived cell class proportions, and the top 5 Principal Components calculated from observed and LD-pruned array genotypes were included as model covariates. To control for potential bias due to the higher variance of lowly expressed genes, we filtered out genes with mean depth <10 reads across 35% of samples and applied apeGLM shrinkage.^42^ Target genes were extracted from the transcriptome-wide results. Differential transcript usage was measured using the tool *DEXSeq*^*43*^ in a multivariate linear model including the same covariates as the differential gene/transcript analysis. Unless otherwise noted all reported *P*-values are adjusted using the Bonferroni method. Gene sets for NF-kB (‘HALLMARK_TNFA_SIGNALING_VIA_NFKB’) and IFN-alpha activation (‘HALLMARK_INTERFERON_ALPHA_RESPONSE’) were taken from the MSigDB database^44^ for expression analysis of genes in molecular pathways related to the 18 target genes.

### Sensitivity analysis of viral load

To assess the impact of viral load on differential gene and transcript expression, a stratified analysis was performed where the subset of participants who exhibited detectable viral load (>200 viral copies/mL of whole blood; N=33) was compared with seronegative control participants (N=361). A parallel comparison was also made between participants who were seropositive but exhibited no detectable viral load (N=194) and the seronegative control participants (N=361).

### Stratified analysis of cocaine use

To assess the impact of cocaine use on gene and transcript expression, a stratified analysis of cocaine use was performed. Participants were grouped based on their use of cocaine (injection cocaine or crack cocaine) in the six months prior to sample collection. Participants who were seropositive for HIV and reported using cocaine, regardless of viral load status (N=121), were compared to seronegative control participants who also reported using cocaine (N=180). A parallel comparison was then made between PLWH who reported no cocaine use (N=106) and seronegative control participants who reported no cocaine use (N=181).

### Drug repurposing analysis

Genes exhibiting significant differential expression were fed into a drug repurposing analysis pipeline^45^ which queries four drug databases: Target Central Resource Database, Open Targets, Therapeutic Target Database, and DrugBank. Drug annotations included known gene targets, and all drugs with gene targets overlapping with the differentially expressed genes in this study were collected and comprehensive cross-resource summaries were collected including clinical status (approved, experimental, preclinical), target selectivity (number of gene targets), and other metrics (e.g., safety).

## RESULTS

### Cohort details

Of the 588 participants in this study, 227 were PLWH (38.6%) with a mean age of 49.6 years (±9.6 SD) at enrollment, and 361 were people not living with HIV with a mean age of 48.9 (±10.2 SD) (**Table 1**). All participants were of genetically predicted European descent, and 74.7% were self-reported male. No significant correlation between age at enrollment and HIV status was observed (*P* = 0.4, univariate logistic regression) nor sex (*P* = 1.0). Opioids were the most used drug (296 individuals, 50%), followed by methamphetamines (203, 34%) and powder cocaine (162, 28%). All PLWH were on ART at the time of blood draw. Viral load was undetectable (<200 viral copies/mL of whole blood) for 194 PLWH individuals. Among the 33 individuals with detectable viral load, the mean viral load was 18,850 viral copies/mL. Focusing on cocaine use (crack and powder), 121 (47%) PLWH reported using cocaine, and 180 (50%) HIV-participants also reported cocaine use.

**Table 1.** Vancouver People Who Inject Drugs Study characteristics.

|  | **HIV<sup>+</sup>**<br>**Non-detectable**<br>**Viral Load**<br>**N=194** | **HIV<sup>+</sup>**<br>**Detectable Viral**<br>**Load**<br>**N=33** | **HIV<sup>-</sup>**<br>**N=361** | **P\*** |
| --- | --- | --- | --- | --- |
| **Age yr. (mean±SD)** | 51.1±8.4 | 48.5±8.4 | 48.9±10.2 | 0.4 |
| **Sex no. (male, %)** | 148 (76.3) | 26 (78.8) | 265 (73.4) | 1.0 |
| **Genetic inferred ancestry no. (EU, %)** | 194 (100) | 33 (100) | 361 (100) | -- |
| **Viral load, copies/mL (mean±SD)** | 37.8±1.3 | 5992.1±8.7 | 0±0 | -- |
| **Cocaine no. (%)** | 58 (29.9) | 9 (27.3) | 95 (26.3) | 1.0 |
| **Crack no. (%)** | 82 (42.3) | 10 (30.3) | 137 (38.0) | 1.0 |
| **Any cocaine no. (%)** | 107 (55.2) | 14 (42.4) | 180 (49.9) | 1.0 |
| **Heroin no. (%)** | 69 (35.6) | 14 (42.4) | 177 (49.0) | 0.03 |
| **Opioid no. (%)** | 86 (44.3) | 15 (45.5) | 195 (54.0) | 0.3 |
| **Meth. no. (%)** | 75 (38.7) | 13 (39.4) | 115 (31.9) | 0.9 |
| **Cannabis no. (%)** | 30 (15.5) | 4 (12.1) | 34 (9.4) | 0.5 |
| **Any stimulant no. (%)** | 151 (77.8) | 22 (66.7) | 242 (67.0) | 0.2 |
| **Any non-prescript no. (%)** | 170 (87.6) | 28 (84.8) | 292 (80.9) | 0.5 |

\*Univariate tests were performed using the full HIV<sup>+</sup> cohort (N=227); P-values were adjusted using Bonferroni method.

### Gene expression differences associated with HIV status

Using negative binomial regression to test for differences in gene expression by HIV status (while adjusting for sex, age, RIN, top 5 genotype PCs, and proportions of 5 immune cell classes), we observed significant differential expression for 9 out of the 18 target genes: *EPSTI1, IFI44L, IFIT3, MX1, NLRC5, PARP9, PLSCR1, RIN2*, and *RSAD2* (**Table 2, Figure 1**), after correcting for multiple comparisons. All 9 of these genes were reported as having proximal hypo-methylated sites in PLWH in prior work and showed upregulated expression in samples from PLWH in the current study.^14-18^

**Table 2.**
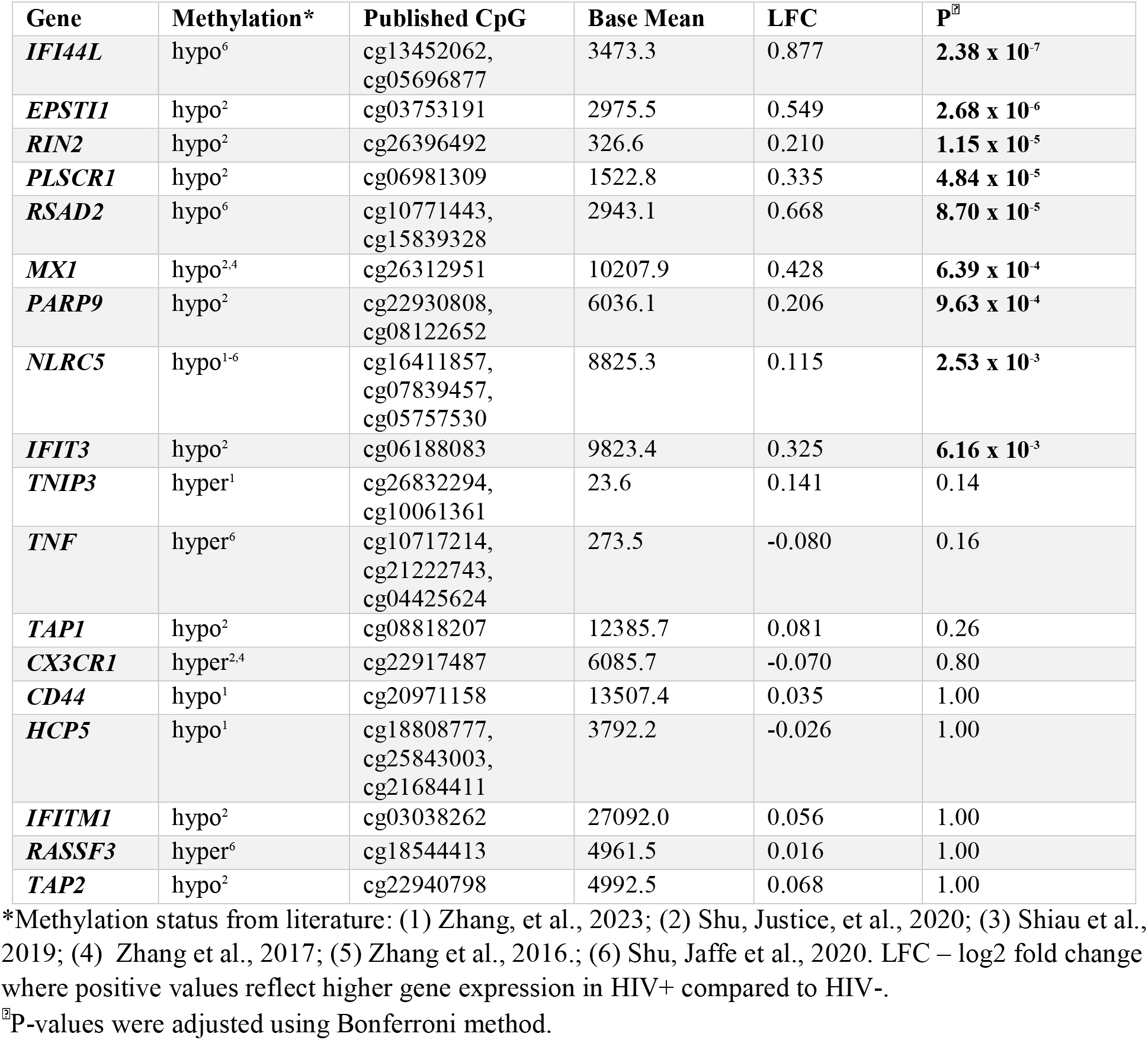
Differential gene expression results for target genes.

**Figure 1.**
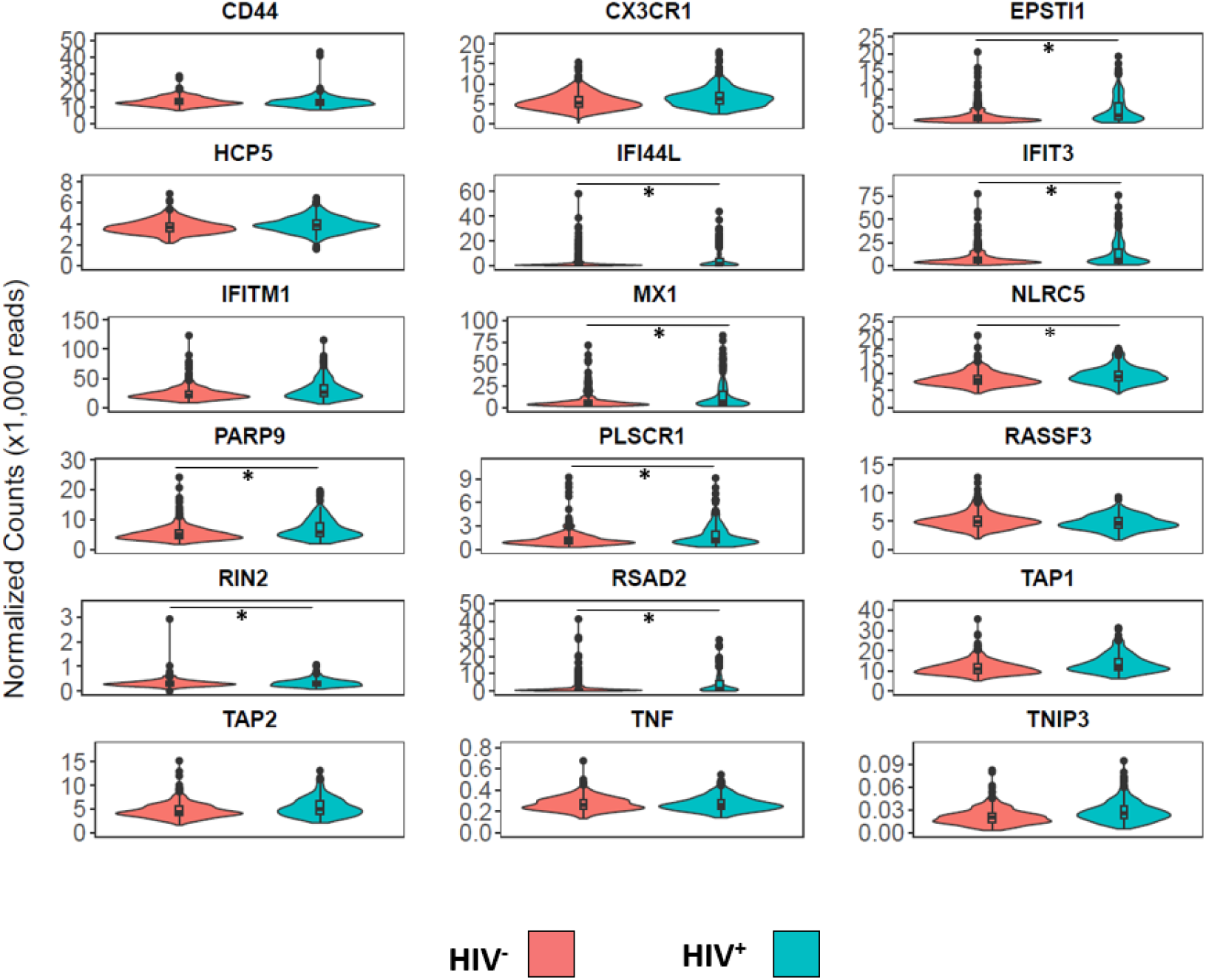
Differential gene expression of 18 targeted genes. Shown are violin plots of normalized mRNA read counts based on HIV status with one gene per panel. Red represents participants who were HIV- and blue/green represents PLWH. \**P* < 0.01

Sensitivity analysis was performed to evaluate the potential influence of HIV viral load on gene expression differences. The full study cohort was stratified into PLWH with detectable viral load (N=33) and those without (N=194), comparing both subgroups to HIV^-^ controls (N=361). Among virally suppressed PLWH, the same nine genes from the full cohort results were again differentially expressed (**Supplemental Table 2**). Among PLWH who had detectable viral load all nine genes plus two additional genes were significantly upregulated: *TAP1* (*P*=0.03) and *TNIP3* (*P*=0.008) (**Supplemental Table 2**). No significant association between normalized transcript counts and viral load was observed.

### Differential transcript expression associated with HIV status

We next examined differential gene isoform expression in the context of HIV status. All nine genes with expression differences at the gene level also showed differential transcript expression. Three additional genes exhibited significantly altered transcript expression, despite showing no differences in overall gene expression. Three transcripts for *CD44* (*ENST00000442151, P <* 0.001; *ENST00000528922, P* = 0.01; *ENST00000531118, P* = 0.04), one transcript for *RASSF3* (ENST00000540088, *P* = 0.02) and two transcripts for *TAP1* (ENST00000487296, *P <* 0.001; ENST00000486332, *P* = 0.003) were upregulated in PLWH relative to HIV^-^ participants **(Figure 2)**. Full differential transcript results are available in **Supplemental Table 3**.

**Figure 2.**
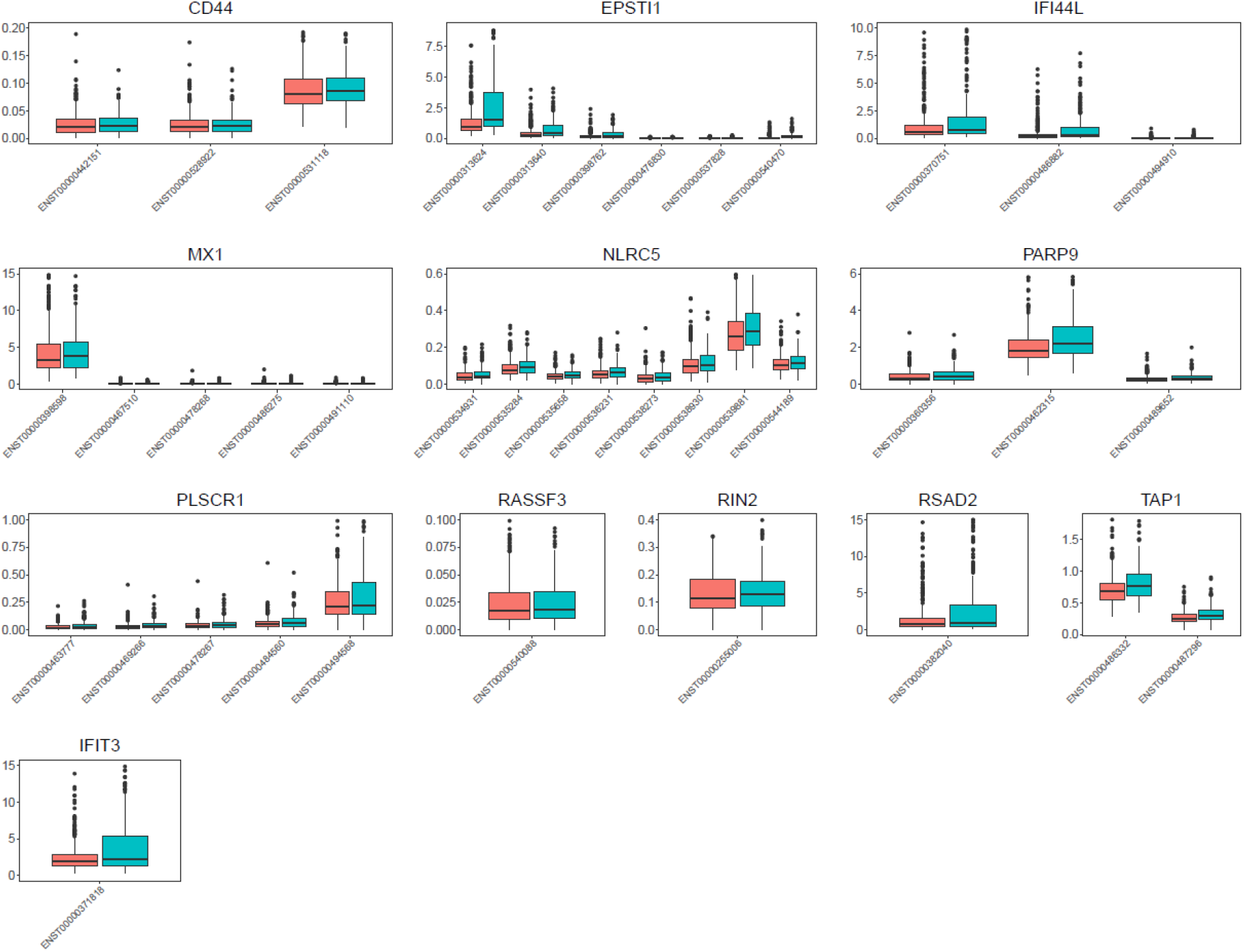
Differential transcript expression results. Shown are boxplots plots of normalized mRNA read counts based on HIV status with one gene per panel. Red represents participants who were HIV- and blue/green represents PLWH. Only transcripts exhibiting *P* < 0.01 are shown.

We next investigated whether these dynamics changed in the context of detectable viral load. Stratification by detectable viral load (N=33) versus undetectable (N=194) compared to seronegative controls (N=361) showed differences in transcript expression dynamics not observed in the full cohort (i.e., regardless of viral suppression status; N=227). Transcripts for genes *CD44, IFIT3, NLRC5, PLSCR1*, and *TAP1* were significantly upregulated in PLWH who had detectable viral load but not in those from PLWH who were virally suppressed (**Supplemental Table 3**). Among these upregulated transcripts, two are protein coding: *ENST00000371811* (*IFIT3)* and ENST00000462666 (*PLSCR1*). Full results of transcript expression analyses stratified by detectability of viral load are available in **Supplemental Table 3**.

### Cocaine Use Stratified Analysis

Given the persistent associations between cocaine use, HIV disease severity and DNA methylation,^15,18^ stratified analysis was performed on cocaine use. Two comparisons were made; (1) recent use of cocaine in PLWH (N=121) versus recent use of cocaine in seronegative individuals (N=180), and (2) no cocaine use in PLWH (N=106) versus no cocaine use in seronegative individuals (N=181). In the context of recent cocaine use, genes *IFI44L* (P=0.003), *MX1* (*P* = 0.03) and *RSAD2* (*P =* 0.01) were significantly upregulated in HIV+ samples but not in the context of non-recent cocaine use (**Supplemental Table 4**). In contrast, one gene, *EPSTI1*, was upregulated in HIV+ samples in the context of non-recent cocaine use relative to HIV-samples (*P* = 0.001), but this upregulation for *EPSTI1* was not observed for recent cocaine use HIV+ samples versus HIV-samples. At the transcript level, two transcripts for gene *PARP9* were significantly upregulated in recent cocaine use HIV+ samples (ENST00000360356, *P* = 0.003; ENST00000489652, *P* = 0.03), as well as one transcript each for genes *MX1* (ENST00000467510, *P* = 0.003), *PLSCR1* (ENST00000469266, *P* = 0.01), and *NLRC5* (ENST00000536231, *P* = 0.008).

### Pathway Enrichment Analysis and Downstream Expression

Pathway analysis was performed on the list of 12 genes exhibiting differential expression either at the gene level or transcript level. Enrichment was observed for the interferon alpha/beta signaling pathway (*P* = 1.75×10^−6^, FDR), and its parent pathways, including interferon signaling (*P =* 1.83×10^−4^, FDR) and Cytokine Signaling in Immune system (*P =* 0.02, FDR; **Table 3**).

**Table 3.** Pathway analysis results for 12 differentially expressed genes.

| Pathway | Genes in Pathway<br>From Study | Total Genes in<br>Pathway | Raw P-value | P-value FDR |
| --- | --- | --- | --- | --- |
| *Interferon alpha/beta signaling* | 7 | 129 | $1.64 \times 10^{-8}$ | **$1.75 \times 10^{-6}$** |
| *Interferon Signaling* | 9 | 397 | $3.45 \times 10^{-6}$ | **$1.83 \times 10^{-4}$** |
| *Cytokine Signaling in Immune system* | 10 | 1099 | $4.73 \times 10^{-4}$ | **0.02** |
| *Immune System* | 11 | 2663 | $4.09 \times 10^{-3}$ | 0.1 |

Next, given that nine of the 18 target genes are themselves affected by interferon-alpha activation, we assessed gene expression changes by HIV status for 80 genes that are upregulated in response to interferon-alpha cytokine,^46^ and 33 of these exhibit significant dysregulation in the current study (*P* < 0.05; **Supplemental Table 6**). *NLRC5*, one of our differentially expressed target genes, transcriptionally activates MHC Class I genes^47^ and regulates the NF-kB pathway.^48^ We tested the hypothesis that downstream targets of *NLRC5* within these pathways would also exhibit differential expression by HIV status. None of the MHC Class I genes were significantly dysregulated in HIV+ samples relative to HIV-samples (**Supplemental Table 6**). However, of the approximately 200 genes identified as targets of NF-kB after stimulation with TNF,^46^ 32 exhibited significant dysregulation by HIV status in our study (**Supplemental Table 6**).

### Drug Repurposing Analysis

The 12 target genes exhibiting expression differences at either the gene or transcript level were next subjected to a drug repurposing analysis to identify any approved or experimental drugs that could be potential therapies (**Supplemental Table 7**). One monoclonal antibody targeting *CD44* was identified: bivatuzumab.

## DISCUSSION

Multiple EWAS have identified and replicated changes in methylation status near multiple genes including *NLRC5, MX1, CX3CR1*, and *TNF* in PLWH compared to people without HIV.^14-19^ No study has assessed whether these methylation changes extend to gene expression changes at these loci until now. We performed a targeted gene expression study in PLWH to evaluate gene expression changes at 18 genes within a combined cohort of people who inject drugs that includes HIV seropositive cases in the VIDUS cohort and seronegative controls from ACCESS. Nine of the 18 target genes were significantly upregulated in HIV+ samples, including the most replicated hypomethylated gene *NLRC5*. Results from this study suggest that a subset of these target genes activate proinflammation and innate immune system pathways in PLWH, especially among those with detectable viral load and PLWH who recently used cocaine.

Hypomethylation at site cg07839457 is observed among PLWH which lies within the promoter region of *NLRC5*.^*14-19*^ This gene is a key regulator of multiple innate immune system pathways, including MHC class I gene expression and NF-kB activation of pro-inflammatory genes.^20,47^ Chronic inflammation and activation of the innate immune system is common in PLWH, leading to increased risk of negative health outcomes such as organ damage, cardiovascular disease, and neurocognitive disorders.^49^ We observed increased expression of *NLRC5* in PLWH, consistent with prior methylation studies. In addition, many targets of NF-kB activation were significantly dysregulated in the current study. However, transcript level analysis revealed a lack of dysregulation attributable to specific protein-coding isoforms. Interestingly, most of the *NLRC5* dysregulation occurred in isoforms annotated with nonsense-mediated decay, retained introns, and processed transcripts without an open reading frame. We hypothesize that excessive transcription of non-coding *NLRC5* isoforms impacts the post-transcriptional activity of protein coding isoforms by potentially regulating translation through lncRNA-mRNA hybridization.^50^ Intron retention may itself be associated with inflammatory response.^51^ Future studies will be required to test the functional impact of *NLRC5* dysregulation in PLWH.

Dysregulation was also observed in the current study for target genes *EPSTI1, IFI44L, IFIT3, MX1, PARP9, PLSCR1, RIN2*, and *RSAD2*. These genes are targets and/or regulators of interferon type I or II signaling,^52-56^ targets of NF-kB activation,^57^ produce proteins with antiviral function,^58^ or activate immune cells.^59^ Prior studies have shown hypomethylation in the region surrounding these genes in PLWH,^14-19^ and the current study confirms the predicted upregulation of gene expression. The upregulation of these genes suggests a systemic dysregulation in proinflammatory signaling via IFN type I and II in PLWH and agrees with prior studies showing chronic inflammation persisting in ART treated chronic HIV.^60^ Supporting this model, 32 NF-kB gene targets and 33 IFN-alpha gene targets were also dysregulated in the current study.

Higher detectable viral load in PLWH is associated with more rapid HIV disease progression.^61^ Some participants in this study exhibited detectable viral load (N=33), and genes *TAP1* and *TNIP3* were additionally upregulated only among this subset. *TAP1* expression is controlled by NLRC5 and helps to traffic antigenic peptides to the cellular membrane for presentation by MHC Class I complex,^62^ and *TAP1* upregulation is observed in infiltrating CD8+ T cells, dendritic cells, and macrophages in many cancer types and outcomes.^63^ Less is known about *TNIP3*, although a recent study found upregulation of *TNIP3* mRNA among TNF-alpha-secreting cells in individuals suffering from major depressive disorder.^64^ Results from the current study are consistent with activated immune function among PLWH with detectable viral load.

The genes *MX1* and *RSAD2* were upregulated among PLWH who reported recent (i.e., last six months) cocaine use compared to seronegative individuals who recently used cocaine, but these genes were not upregulated in the comparison of PLWH reporting no cocaine use versus seronegative individuals with no cocaine use. Similarly, upregulation was observed among transcripts for genes *PARP9, PLSCR1, NLRC5*, and *MX1* in PLWH who used cocaine recently versus seronegative participants who used cocaine recently, but this was not the case in the comparison of PLWH reporting no cocaine use versus seronegative participants reporting no cocaine use. Prior work has shown methylation changes mediating the association between cocaine use and HIV disease severity,^18^ and gene expression results in the current study support this model.

Drug repurposing analysis revealed one of the 12 genes in this study, *CD44*, exhibiting dysregulated expression had one drug known to target it: bivatuzumab mertansine.^65^ This prodrug was developed Three isoforms of *CD44*, including one protein-coding isoform ENST00000442151, were significantly upregulated in the current study. Thus, use of bivatuzumab may be considered in the context of HIV disease progression.

This study comes with limitations. First, this was an observational cohort study of a single time point, preventing dissection of the timing of gene dysregulation. Future longitudinal studies will help to elucidate the dysregulation cascade for these genes. Stratified analysis of PLWH with detectable viral load was limited in power (N=33). Cocaine use was measured via self-reported questionnaire about the six months prior to study enrollment, and responses were binary. Future studies should consider measuring the frequency and/or intensity of cocaine use leading up to sample collection to assess whether these aspects drive gene expression dysregulation even further.

In conclusion, we have confirmed the gene expression dynamics predicted from prior methylation studies of HIV acquisition and disease progression.^14-19^ *NLRC5* and eight other target genes were upregulated in PLWH. Results from this study support a model of epigenetic changes resulting in systemic gene dysregulation within proinflammation and innate immune system pathways in PLWH. Drugs targeting *CD44* gene activity may help to alleviate these inflammatory effects and potentially influence HIV-related comorbidities.

## Data Availability

All data produced in the present study are available upon reasonable request to the authors

